# Predicting bloodstream infection outcome using machine learning

**DOI:** 10.1101/2021.05.18.21257369

**Authors:** Yazeed Zoabi, Orli Kehat, Dan Lahav, Ahuva Weiss-Meilik, Amos Adler, Noam Shomron

## Abstract

Bloodstream infections (BSI) are a main cause of infectious disease morbidity and mortality world-wide. Early prediction of patients at high risk of poor outcomes of BSI is important for earlier decision making and effective patient stratification. We developed electronic medical record-based machine learning models that predict patient outcomes of BSI. The area under the receiver-operating characteristics curve was 0.82 for a full featured inclusive model, and 0.81 for a compact model using only 25 features. Our models were trained, using electronic medical records that include demographics, blood tests, and the medical and diagnosis history of 7,889 hospitalized patients diagnosed with BSI. Among the implications of this work is implementation of the models as a basis for selective rapid microbiological identification, toward earlier administration of appropriate antibiotic therapy. Additionally, our models may help reduce the development of BSI and its associated adverse health outcomes and complications.

## 1 Introduction

Bloodstream infections (BSI) can lead to prolonged hospital stays, and life-threatening and aggressive complications, in addition to high costs to health care systems [1–4]. Increasing rates of antimicrobial-resistant pathogens, particularly gram-negative bacteria, limit treatment options; this often prompts empirical use of broad-range antibiotics [5]. Therefore, timely and critical assessment of available microbiology results are necessary to ensure that individuals with BSI receive prompt, effective, and targeted treatment, for optimal clinical outcomes [5]. However, the current standard-of-care, which mostly depends on blood culture-based diagnosis, is often extremely slow [6].

Sepsis is a life-threatening medical condition, defined as the body’s systemic immunological response to an infectious process, which may cause end-stage organ dysfunction and eventually death [7]. Several studies have utilized electronic medical records (EMRs) to construct prediction models for mortality from sepsis [8, 9] and sepsis onset [12–15]. From a clinical perspective, these types of early warning systems may be useful in detecting patients at risk of BSI, and typically provide information at a certain point in time (for example, preoperatively [16]) or in a certain time-window before deterioration (see review in [17]). Identifying a patient at risk can trigger early goal-directed therapy regarding confirmation of infection, administration of antimicrobial therapy, and transition to the intensive care unit (ICU).

In contrast to studies that aimed to detect patients at risk of BSI, the current study focused on patients with confirmed infections. Based on EMRs of patients hospitalized with positive blood cultures, we constructed machine learning models that predict poor outcomes of hospitalized patients with BSI (Fig. 1; see Methods). Our prediction occurs at a certain point of time: just after the identification of bacterial morphology on direct gram stain from a positive blood culture, yet well before performing specific pathogen identification. At this point of time, early goal-directed clinical practice can be diverted to patients at risk for poor clinical outcomes. In addition, the prediction can facilitate deciding at the microbiology lab, between using the traditional lengthy pathogen identification routine or rapid microbiological identification techniques [18]. A forewarning system could direct the required resources to the patients with BSI in the greatest need.

**Figure 1.**
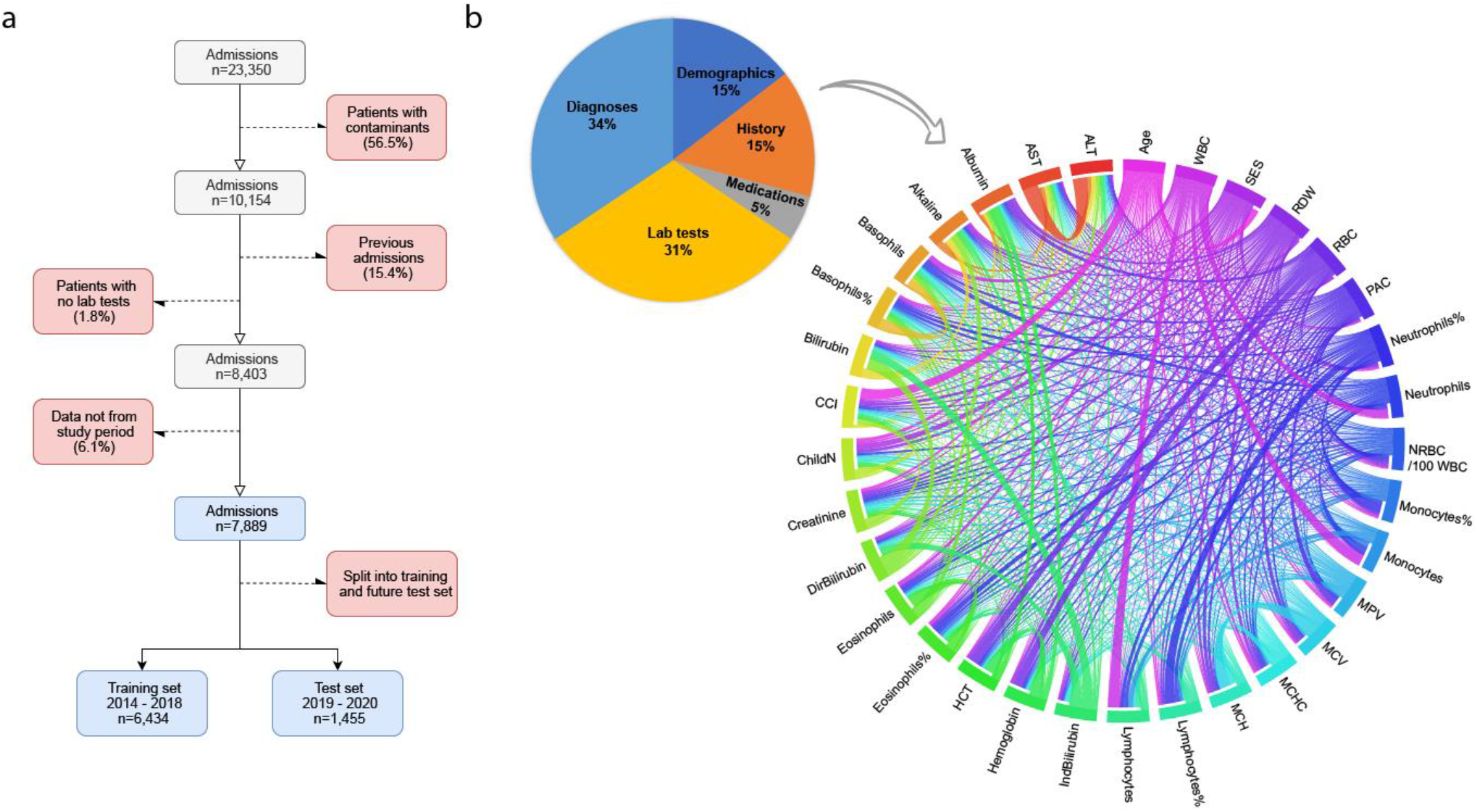
Data and cohort characteristics. **a**, Cohort selection. Bloodstream infection was identified as a positive culture test. Next, patients with only contaminants were excluded (a list of bacteria classified as contaminants is available in the supplementary information). Subsequently, previous admissions for each patient and patients with no lab tests information were excluded. Finally, the cohort was divided into training and validation sets (see Methods). **b**, Feature modality distribution. Pie charts are divided according to the sum of data points in each feature set. A substantial proportion of the data originates from laboratory test results during current or previous admissions. The Circos plot shows the correlation between continuous features from the entire cohort (test and training sets). Correlation strength is determined by Pearson correlation, thicker bands correspond to a stronger Pearson correlation coefficient.

## 2 Results

### The inclusive model

The characteristics of the population used for the training and testing of the inclusive model are described in Table 1 (see Methods). From 7,889 adults with a BSI, 2,590 (32.8%) positive composite outcomes were recorded. The contribution of each feature of the inclusive model to predict the composite outcome was measured by SHaply Additive exPlanations (SHAP) scores, for each patient (Fig. 2). The predictive contribution of missingness (gray points) was also assessed. Accordingly, a missing value of a feature (e.g., albumin) serves as a signal regarding the patient’s risk. SHAP values of three selected variables – age, monocyte %, albumin – are presented in Fig. 2b-d. The top 20 features ranked by the SHAP scores were also calculated (see Supplementary Fig. 1). The calibrated model was tested on a subset of the future test set, which comprised only patients in the ICU (the results are presented in Supplementary Fig. 2).

**Table 1.** Population Characteristics. CCI = Charlson Comorbidity Index.

|  |  | Training set | Test set |
| --- | --- | --- | --- |
| **General** | **N** | 6434 | 1455 |
|  | **Age [median (±)]** | 74 (24) | 73 (22) |
|  | **Females [%]** | 47.7 | 46.25 |
| **CCI** | **Mild [%]** | 20.66 | 20.03 |
|  | **Moderate [%]** | 40.4 | 37.94 |
|  | **Severe [%]** | 38.94 | 42.03 |
| **Admission type** | **Elective [%]** | 5.08 | 5.54 |
|  | **Emergency [%]** | 91.17 | 90.36 |
|  | **Urgent [%]** | 3.75 | 4.1 |
| **Other** | **Infectious background [%]** | 9.71 | 9.42 |
|  | **Surgery during hospital stay [%]** | 14.45 | 13.33 |
| **Clinical outcomes** | **Short-term mortality [%]** | 23.17 | 18.49 |
|  | **Long hospital stay [%]** | 12.17 | 9.07 |
|  | **Mechanical ventilation [%]** | 1.23 | 0.97 |
|  | **Composite score [%]** | 34.24 | 26.6 |

**Figure 2:**
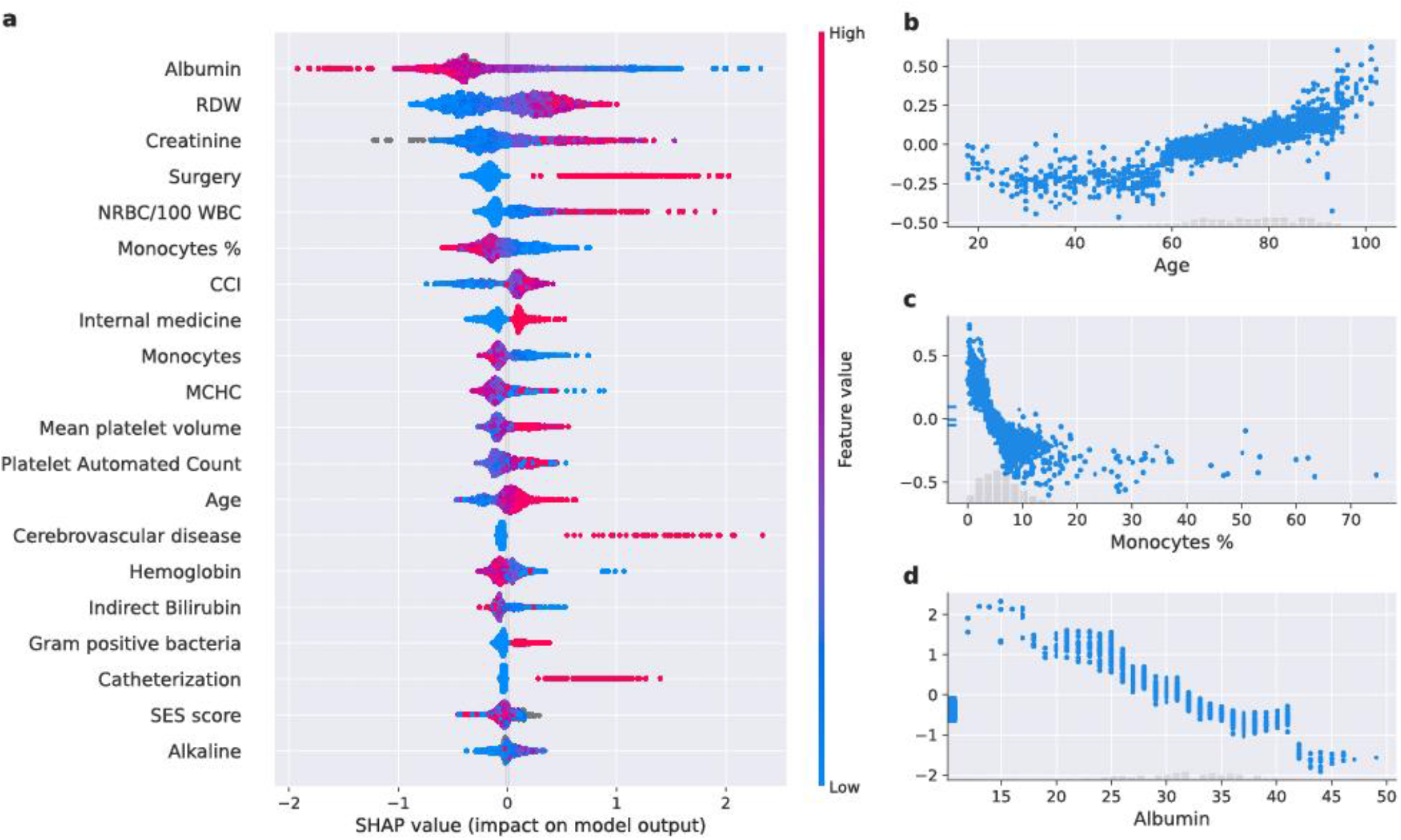
Feature analysis of the inclusive model. **a**. A summary plot of the SHaply Additive exPlanations (SHAP) values for each feature. From top to bottom, the features are ordered by their overall influence on the final prediction (sum of SHAP values). In each feature (line), each point represents a specific case (individual), with colors ranging from red (high values of the predictor) to blue (low values of the predictor). Gray points signal missing values. A point’s location on the X-axis represents the SHAP value—the effect the variable had on the prediction in a given individual; points further right indicate greater risk, and points to the left indicate lesser risk. The vertical line in the middle represents no change in risk. **b** A plot of SHAP for different values of age (years). The light histogram along the X-axis shows the density of the data. **c** A plot of SHAP for different values of monocytes percentage (%) in the blood. The light histogram along the X-axis shows the density of the data. **d** A plot of SHAP for different values of albumin (g/L). The light histogram along the x-axis shows the density of the data. **a-d** are based on the future test set, n = 1,455 unique patients.

Performance of the inclusive prediction model on the test set showed area under the receiver-operating characteristics curve (auROC) of 0.82 (95% confidence intervals (CI): 0.80–0.845), which indicates good discrimination; and an area under the precision-recall curve (auPRC) of 0.65 (95% CI: 0.61– 0.70) (Fig. 3). The calibration plot, which runs very close to the diagonal, shows excellent calibration (Fig. 3c).

**Figure 3:**
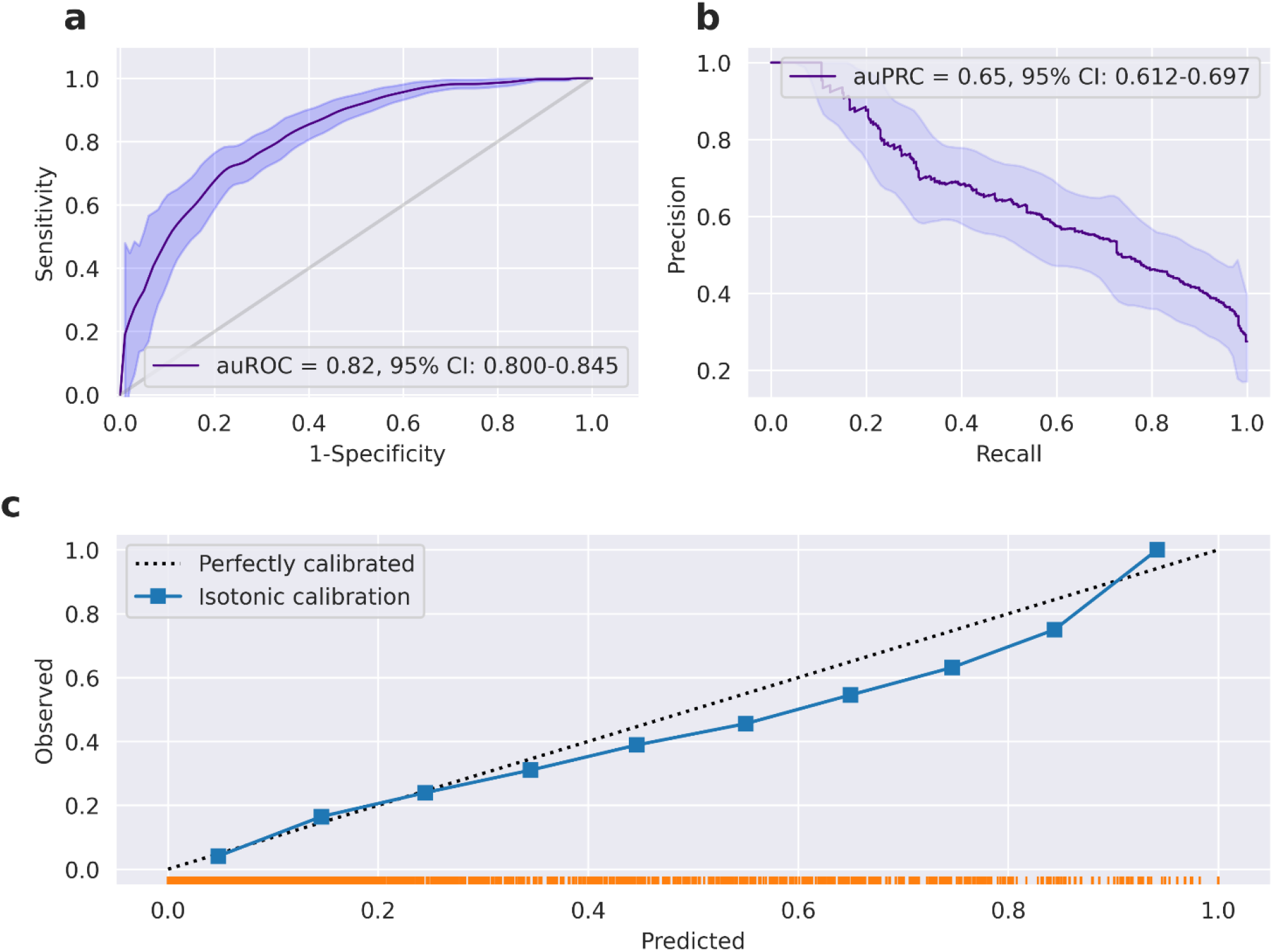
Performance of the inclusive model. **a**. Receiver-operating characteristics (ROC) curves for predictions of the inclusive model on the prospective test set. The light band around the curve represents pointwise 95% confidence intervals derived by bootstrapping. **b**. A plot of the precision (positive predictive value, PPV) against the recall (sensitivity) of the predictor for different thresholds. The light band around the curve represents pointwise 95% confidence intervals derived by bootstrapping. **c**. Calibration plot, plotting the observed outcome against the predicted probabilities. The diagonal gray line represents perfect calibration. A smoothed line is fit to the curve, and points are drawn to represent the averages in ten discretized bins. The rug under the plot illustrates the distribution of predictions.

### The compact model

The numerous EMR features, more than 600, incorporated in the inclusive model render its external validation and the reproducibility of results very difficult. This limits its applicability to other hospitals. Hence, we established a simpler and more compact prediction model that incorporates the features with the greatest impact on outcome, and that are most commonly listed in EMR datasets from other hospitals. To this end, we trained and evaluated the performance of a compact model with only 25 features, including simple demographic information, a single blood test, and a brief medical history (Fig. 4.d). We used the same cohort of patients of the training and test sets from the previous analysis (see the inclusive model). The compact model achieves an auROC of 0.81 and auPRC of 0.63 (Fig. 4.a and Fig. 4.b), which is only slightly lower than the values of the inclusive model, with good calibration (Fig. 4.c). SHAP scores are shown in Figure 5.

**Figure 4:**
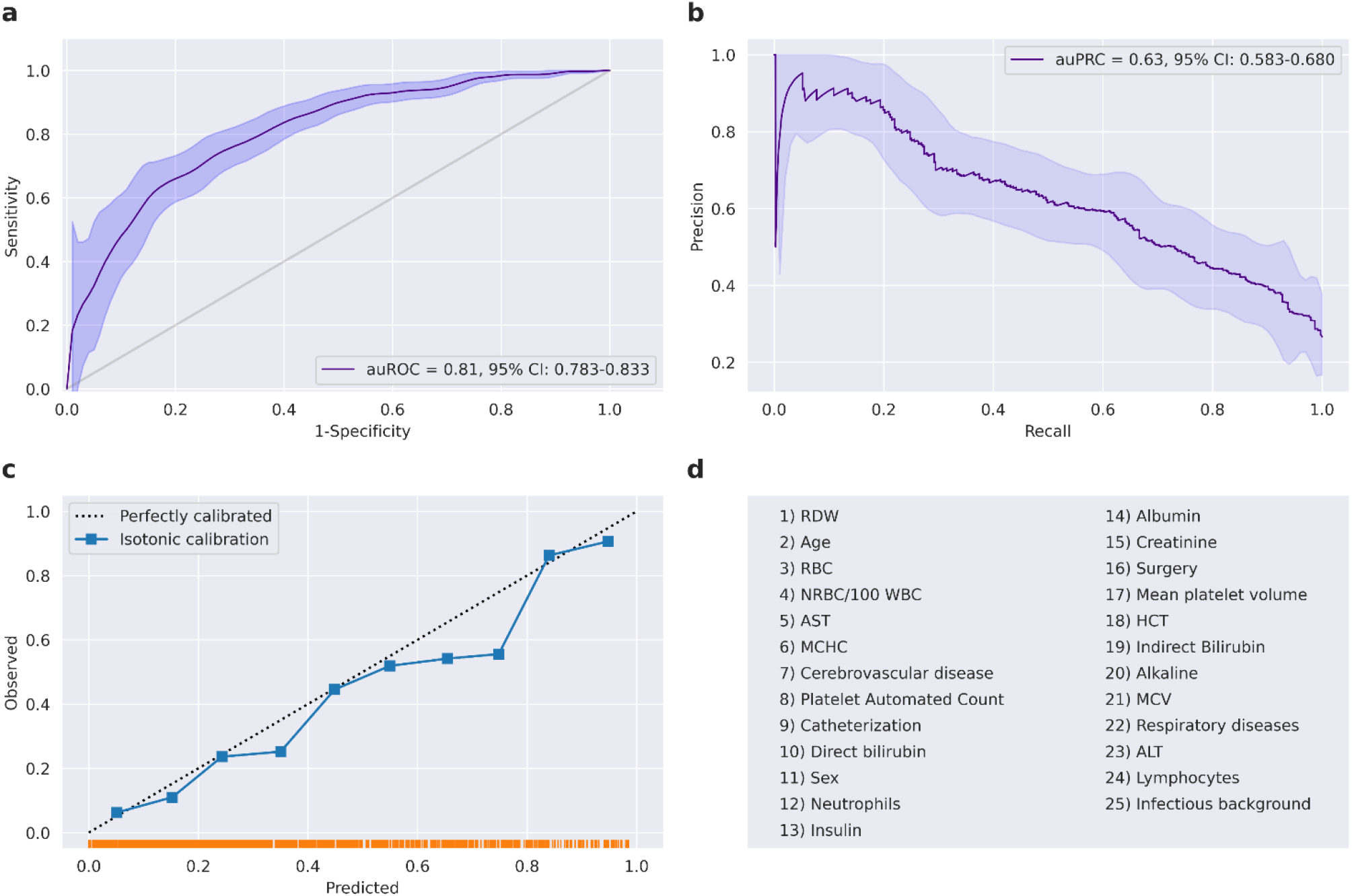
Performance of the compact model. **a**. Receiver-operating characteristics (ROC) curves for predictions of the compact model on the prospective test set. The light band around the curve represents pointwise 95% confidence intervals derived by bootstrapping. **b**. A plot of the precision (positive predictive value, PPV) against the recall (sensitivity) of the predictor for different thresholds. The light band around the curve represents pointwise 95% confidence intervals derived by bootstrapping. **c**. Calibration plot, plotting the observed outcome against the predicted probabilities. The diagonal gray line represents perfect calibration. A smoothed line is fit to the curve, and points are drawn to represent the averages in ten discretized bins. The rug under the plot illustrates the distribution of predictions. **d**. All 25 features used by the compact model.

**Figure 5:**
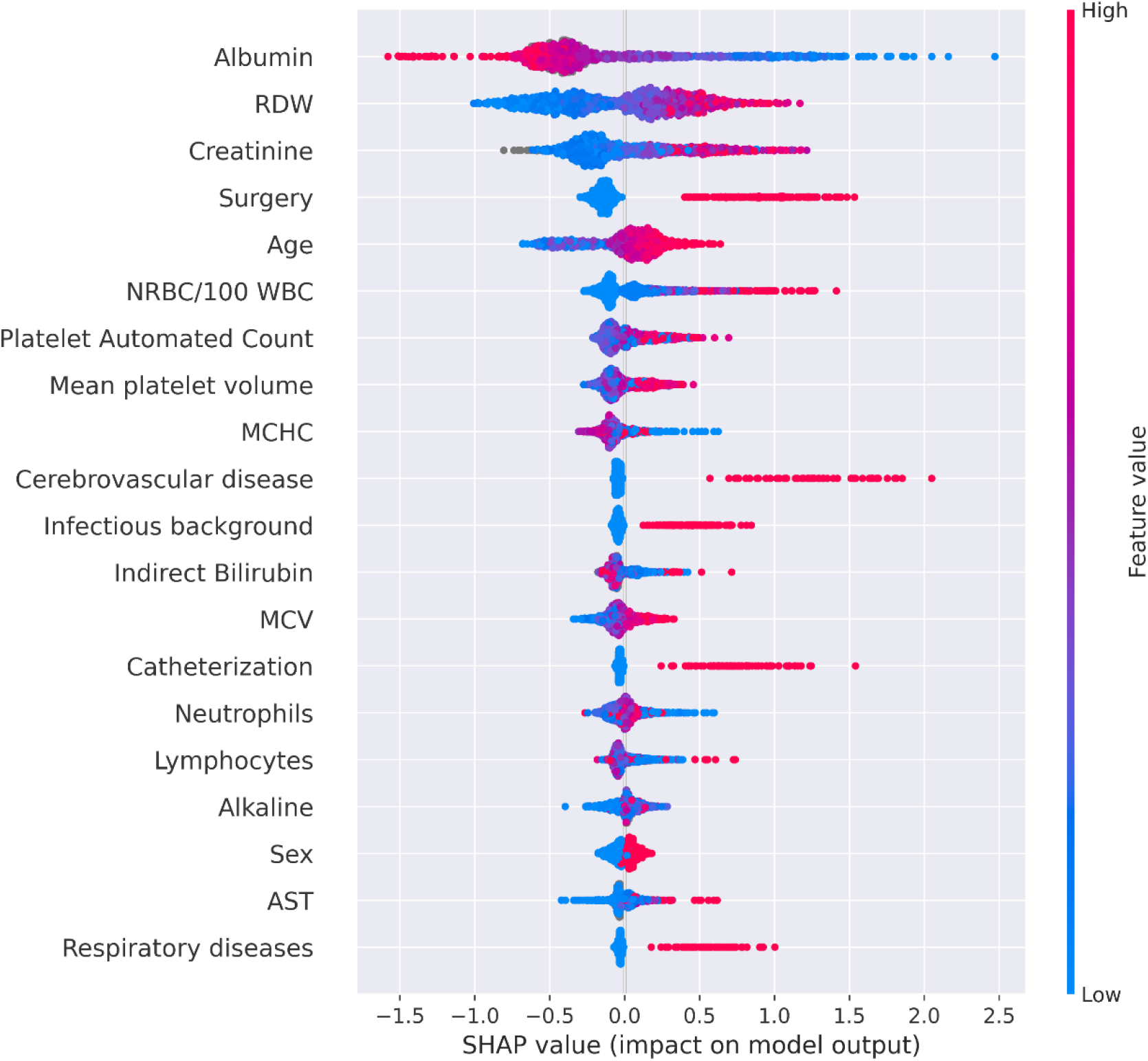
A summary plot of the SHAP values for each feature of the compact model. From top to bottom, features are ordered by their overall influence on the final prediction (sum of SHAP values). In each feature (line), each point represents a specific case (individual), with colors ranging from red (high values of the predictor) to blue (low values of the predictor). Gray points signal missing values. A point’s location on the X-axis represents the SHAP value—the effect the variable had on the prediction in a given individual. The points further to the right indicate that for the given individual, the covariate contributed to increasing the risk. Points to the left indicate that the covariate contributed to decreasing the risk. The vertical line in the middle represents no change in risk. Values for the feature ‘Sex’ are 0 for female, and 1 for male.

The code to test this model is available in our GitHub repository.

## 3 Discussion

In this study, we examined the ability to utilize EMRs to predict a composite poor outcome of patients with BSI, which may promote both rapid interventions and patient stratification. Several scoring systems have been developed in recent years for stratifying the risk of patients with sepsis [8, 12–15, 17], but not for outcomes of patients with BSI. In addition, none are commonly used in routine practice nor recommended according to current guidelines.

Our results show that EMRs can be used to produce accurate predictions of BSI outcomes. Our retrospective analysis demonstrates the feasibility of accurate prediction of BSI outcomes using data available in EMRs. In the inclusive model, the prediction was after a gram-stain, and yielded an auROC of 0.82, and an auPRC of 0.65. In the compact model, the prediction was based on only 25 features available at the time of culture, not including gram-stain results; and yielded an auROC of 0.81, and an auPRC of 0.63. In addition to the well-established risk factors for complications from BSI, such as age and previous infections, our analysis revealed less-known factors as highly predictive of a poor outcome from BSIs. The main factors that were identified as increasing risk included: red cell distribution width, albumin, and creatinine. Red cell distribution width and albumin have been associated with mortality, and have been used as prognostic markers in a number of studies [19–26]. The Charlson Comorbidity Index has also been found to predict mortality in various medical situations [27, 28]. However, other factors revealed as central by our analysis, such as serum creatinine values and monocyte counts, are less well recognized and used as predictors of a poor outcome in patients with BSI.

Although maximal model explainability requires using the patient’s entire EMR, we demonstrated that a subset of features, available from only simple demographic information and a single blood test, enables accurate prediction with only a slight decrease in auROC, from 0.82 in the inclusive model to 0.81 in the compact model. This may enable accurate BSI outcome estimation by embedded systems in emergency departments of hospitals.

Our work has several clinical applications. For instance, it can help select individuals at high-risk for BSI, for whom hospitalization in the ICU, treatment with broad-spectrum antibiotics, or effective early-stage rapid microbiological identification should be considered.

The benefit of early microbiological identification for patient outcomes has been well described in the medical literature [5, 29] and the *time-to-administration* of appropriate antibiotics to treat BSI is an important predictor of outcomes [30]. The current study therefore paves the route for future randomized control trials to further study the effectiveness of implementing a model for early prediction of BSI outcomes, possible preventive interventions, and more efficient selection of patients for advanced microbiological diagnostic testing, thus reducing the time-to-administration of appropriate antibiotics.

Our study has several limitations. First, our prediction model is based on retrospective EMR data, which have inherent biases and are influenced by the interaction of the patient with the health system [31]. However, these biases are partially mitigated in this study, since the data contain information originating from a public hospital serving a very large population, and since the outcome of the model is based on information that is accurately and comprehensively documented in the EMRs. Another limitation of the study is that we assessed BSI outcomes only in patients who were already diagnosed with BSI (having a positive blood culture). This makes it more difficult to generalize to the entire emergency room population. Finally, the predictor was trained and validated on EMRs from Tel Aviv Sourasky Medical Center (TASMC), composed of patients from and around Tel Aviv, Israel. Nonetheless, TASMC is a public hospital, and the medical system in Israel is accessible to the entire population.

Applicability of the model to other hospital populations needs to be shown. However, the large size of the data, and the comprehensive validation process and its result, namely, validation of the utility of established risk factors for a poor BSI outcome, all support the model’s generalizability to other hospitals. Given the additional complexities introduced by the machine learning algorithms, we sought to ensure that sufficient information would be provided to enable our model to undergo external validation [32, 33]. This drove us to develop the compact model, which is based on the features with the greatest influence on the overall prediction, and that are easily accessible in EMR datasets from other hospitals. This compact model achieved only a slightly reduced auROC of 0.81. We made the compact model available in our GitHub repository (see code availability) and we encourage researchers with similar data from other hospitals to test it.

In conclusion, our work demonstrates that accurate and calibrated predictions of BSI outcomes early in a hospital admission can be achieved. Earlier and better characterization of patients with BSI could potentially reduce the development of BSI and its associated adverse health outcomes and complications. Our predictive model could become the basis of selective, rapid microbiological identification, and also contribute to various decisions such as ICU hospitalization and administration of broad-spectrum antibiotics. Future prospective studies, as well as those on populations from other hospitals, are needed to evaluate the clinical impact of the model.

## 4 Methods

### Study design, population, and definition of outcome

The study was designed as a retrospective cohort study for the development and validation of a clinical prediction model. The study was performed at the TASMC, a 1,500-bed public tertiary care center, and the only general hospital serving the population of Tel-Aviv, the most populous city in Israel, of all socioeconomic backgrounds. Data processing, model training and analyses were performed at the TASMC Data Science Department and the Faculty of Medicine at Tel Aviv University.

The study included EMRs of adults hospitalized with a positive blood culture (bacterial only) in the period between January 2014 and January 2020. The year 2014 was determined as the starting point since frequent changes in variable identifiers occurred in the preceding years. Patients’ EMRs included demographics, laboratory test results, previous diagnoses recorded at TASMC, recorded medical history, and initial gram-stain morphology of positive blood cultures that are reported by phone.

The models were developed according to features extracted from various modalities available in EMRs of patients hospitalized with a positive blood culture at the TASMC, and predicted a composite poor outcome, defined as **at least one** of the following:

- Short term in-hospital mortality within 10 days of a culture.
- Mechanical ventilation in the 10 days after the culture.
- Prolonged length of stay (>6 weeks).

The study flow chart is presented in Fig. 1. Exclusion criteria were: the absence of laboratory data of medical history information, age younger than 18 years or older than 100 years, and patients’ explicit objection to the use of their medical data for research purposes. Before initiating any analysis, the study population was divided into a training set that included 6,434 admissions (from years 2014-2018) and a validation set that included 1,455 admissions during 2019 and the first month of 2020. The model was also validated on two subsets of the validation set, which comprised only patients from ICUs or only patients from the emergency room. Both these subsets posed a high challenge to the model regarding its generalization. The prospective validation cross-sections were performed to emulate the model’s use in practice and in real world situations.

### Variable and feature selection

To evaluate whether EMR-derived information might accurately predict outcomes of patients with BSI, we compiled a set of 606 features. All these features were available at the time a blood sample was sent for culture, except for the gram-stain information (used only in the inclusive model). With all these features, we trained a gradient-boosting model, the inclusive model, to predict the probability that each held-out sample (patients not included in the training set) would have a poor outcome. Distributions of the various modalities of the features used are depicted in Fig. 1b. In addition, we trained and tested a compact model, comprising 25 features, for application on EMRs from other hospitals.

Each of the 606 features was assigned a category. For more comprehensive representation, some features within a category were combined, such that all the features could be represented using 96 variables. These variables were used to create the pie chart in Fig 1.b. The following list describes the mechanism for generating the features, and for grouping them:

A. Demographics (238 features, 14 variables when grouped)
  - Contains features such as age, sex, and number of children. A total of 228 categorical features, which included birth country and nationality, were grouped to five variables.
B. History (108 features, 14 when grouped)
  - Contains medical history that is not documented as diagnosis history. This includes unit information (ICU, emergency room, etc.), surgery information, and chest pain. A total of 97 categorical features were grouped to three variables.
C. Medications (five features)
  - Contains binary information about five medications: general diabetes drugs, insulin, anti-coagulants, anti-aggregants, and coumadin on admission.
D. Laboratory (33 features, 30 when grouped)
  - Contains laboratory test results. Four categorical features, describing gram staining results, were combined into one variable. All other features in this category had continuous numeric values.
E. Diagnoses (222 features, 33 when grouped)
  - Diagnoses history is recorded in TASMC as ICD-9 codes. The ICD-9 code hierarchy was used to group these features into 33 variables represented by ICD-9 codes of higher hierarchy.

### Analysis platform

All computational analyses were performed on a secure compute cluster environment located at TASMC. Python 3, with numpy, pandas, and scikit-learn formed the backbone of the data-processing pipeline.

### Development of the models

Predictions were generated using a gradient-boosting machine model built with decision-tree base-learners [34]. Such models have demonstrated efficacy in prediction, using tabular data [35], and have been incorporated in several successful algorithms in the field of machine learning [36]. We implemented the gradient-boosting predictor trained with the LightGBM [37] Python package. LightGBM has shown effectiveness on clinical and patient tabular data in particular, and was adopted by many recently published models [38–43]. Missing values were inherently handled by the LightGBM predictor [37, 44, 45]. The validation set was used for early stopping [46], with auROC as the performance measure. The hyperparameters used are available at our GitHub repository. Two LightGBM classifiers with differing complexity were developed: the inclusive and compact models. The compact model is available at our GitHub repository (see Code Availability).

### The mechanistic basis of the models

To identify the principal features driving model prediction, SHAP values [47] were calculated. These values are suited for complex models such as artificial neural networks and gradient-boosting machines [48]. Originating in game theory, SHAP values partition the prediction result of every sample into the contribution of each constituent feature value. This is done by estimating differences between models with subsets of the feature space. By averaging across samples, SHAP values estimate the contribution of each feature to overall model predictions. A higher value indicates that a feature has a larger impact on the model, which indicates that the feature is more important.

### Calibration of the models

We analyzed the calibration (observed risk versus raw prediction score) of our proposed inclusive and compact models. The raw prediction scores produced by the machine learning model (LightGBM) were calibrated and evaluated on the test-set. We used isotonic regression, which fits a rank-preserving transformation between the original scores and transformed scores; and minimizes the deviation between the target label and the prediction score. We used the scikit-learn library (version 0.20.0) for fitting the isotonic regression model. Ten prediction score bins were used, with regular spacing between the minimum/maximum prediction score produced by a model. We noticed that the raw scores were already well-calibrated. Curves for isotonic and sigmoid calibrations, as well as for the raw model are presented in Figure 6.

**Figure 6.**
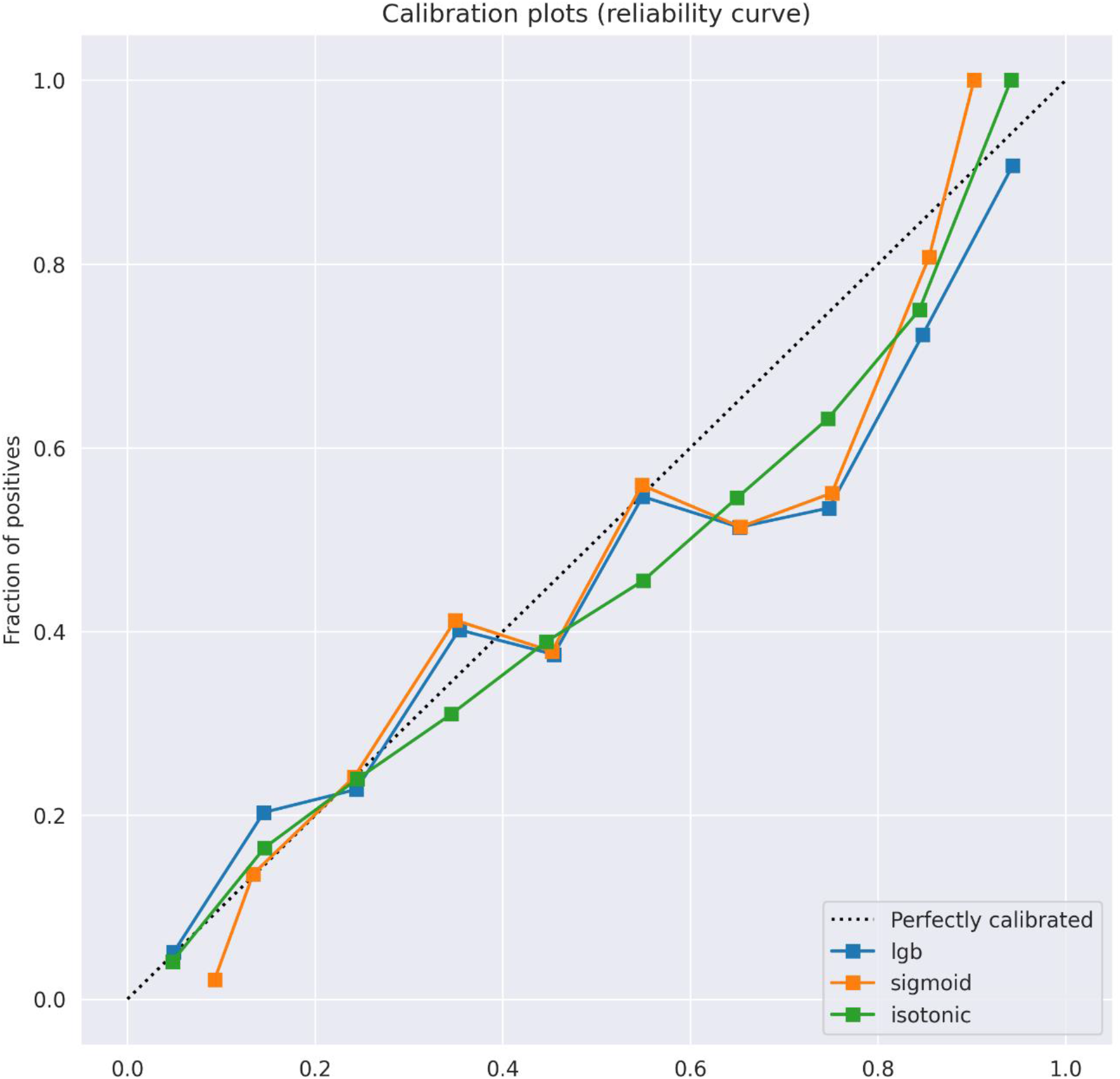
Calibration plots of the observed outcome against the predicted probabilities. The diagonal gray line represents perfect calibration. A smoothed line is fit to the curve, and points are drawn to represent the mean values in ten discretized bins. Blue, orange, and green lines correspond to the original uncalibrated model, the model after sigmoid calibration, and after isotonic calibration, respectively.

### Evaluation of the models

The models were scored on the test set using the auROC. In addition, plots of positive predictive value (PPV) against sensitivity (precision-recall curve) were drawn across different thresholds. For all the thresholds from all the ROC curves, metrics were calculated, including sensitivity, specificity, PPV, negative predictive value, false positive rate, false negative rate, false discovery rate, and overall accuracy (Supplementary Dataset 1). Confidence intervals for the various performance measures were derived through resampling, using the bootstrap percentile method [49] with 1,000 repetitions.

### Ethics declarations

This study (TLV-0684-18) was approved by the TASMC Institutional Review Board (IRB). All methods were performed in accordance with the IRB policy, guidelines, and regulations. Informed consent was waived by the IRB, as all identifying details of the participants were removed before any computational analysis.

## Data Availability

The data that support the findings of this study are from TASMC. Access restrictions apply to these data and they are therefore not publicly available. Due to these restrictions, these data can be ac-cessed only by request to TASMC or the authors.

## 5 Data Availability statement

The data that support the findings of this study are from TASMC. Access restrictions apply to these data and they are therefore not publicly available. Due to these restrictions, these data can be accessed only by request to TASMC or the authors.

## 6 Code Availability statement

Hyperparameters for the models and the analytic code of the compact model are available at: https://github.com/nshomron/infecpred.

## Acknowledgements

Y.Z. and D.L. are partially supported by the Edmond J. Safra Center for Bioinformatics at Tel-Aviv University. The Shomron lab is partially supported by the Adelis Foundation.

## Author Contribution

Y.Z., O.K., A.WM., A.A., N.S. designed the study and wrote the paper. Y.Z. developed the models.

O.K. collected the data. Y.Z. and O.K. did the statistical analysis. D.L. contributed to the development of the compact model.

