## Supplementary figures for "Predicting bloodstream infection outcome using machine learning"

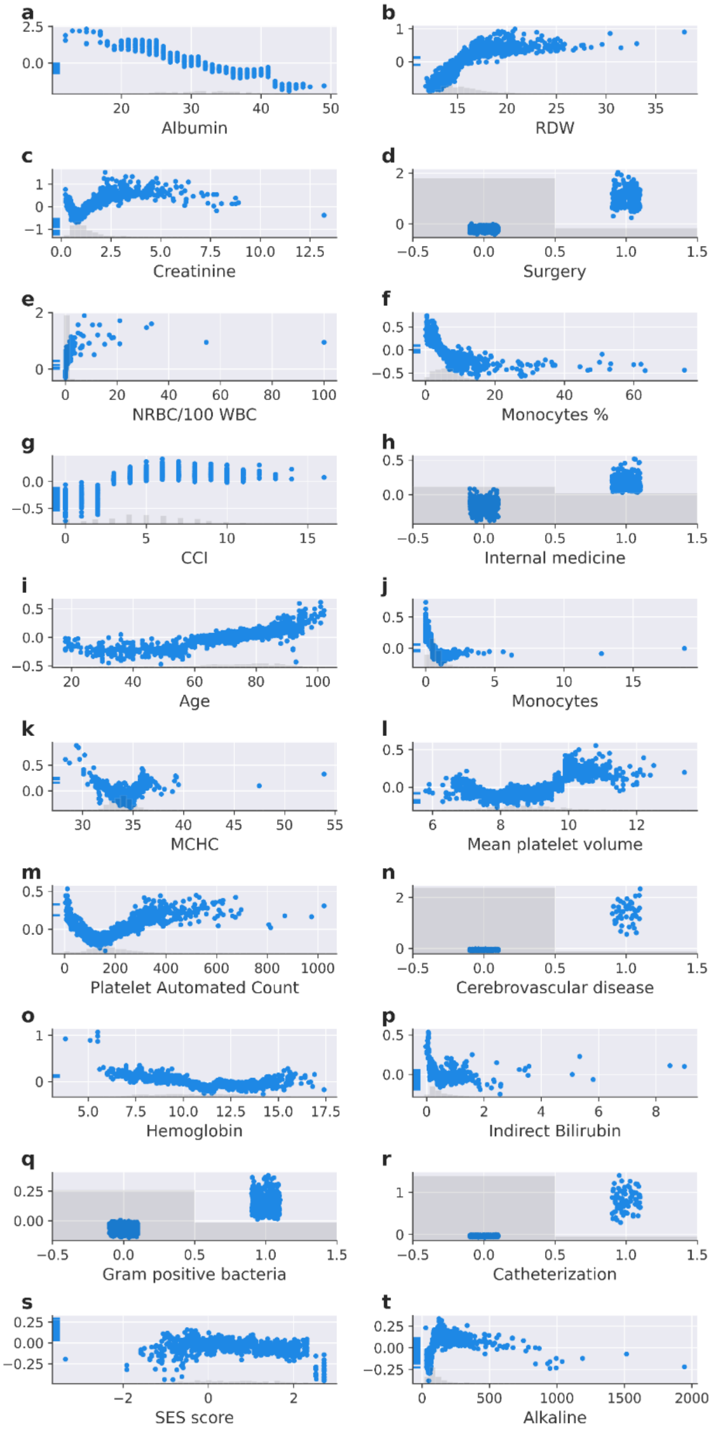


**Supplementary Figure 1. a-t.** Scatter plots of SHAP for the different values of the top 20 features for the inclusive model. The light histogram along the X-axis shows the density of the data. Albumin (g/L), RDW (%), Creatinine (mg/dL), NRBC/100 WBC (%), Monocytes (%), Age (years), Monocytes (10e3/ML), MCHC (g/dL), Mean platelet volume (fL), Platelet automated count (10e3/ML), Hemoglobin (g/dL), AST (U/L), indirect bilirubin (mg/dL), alkaline (U/L).

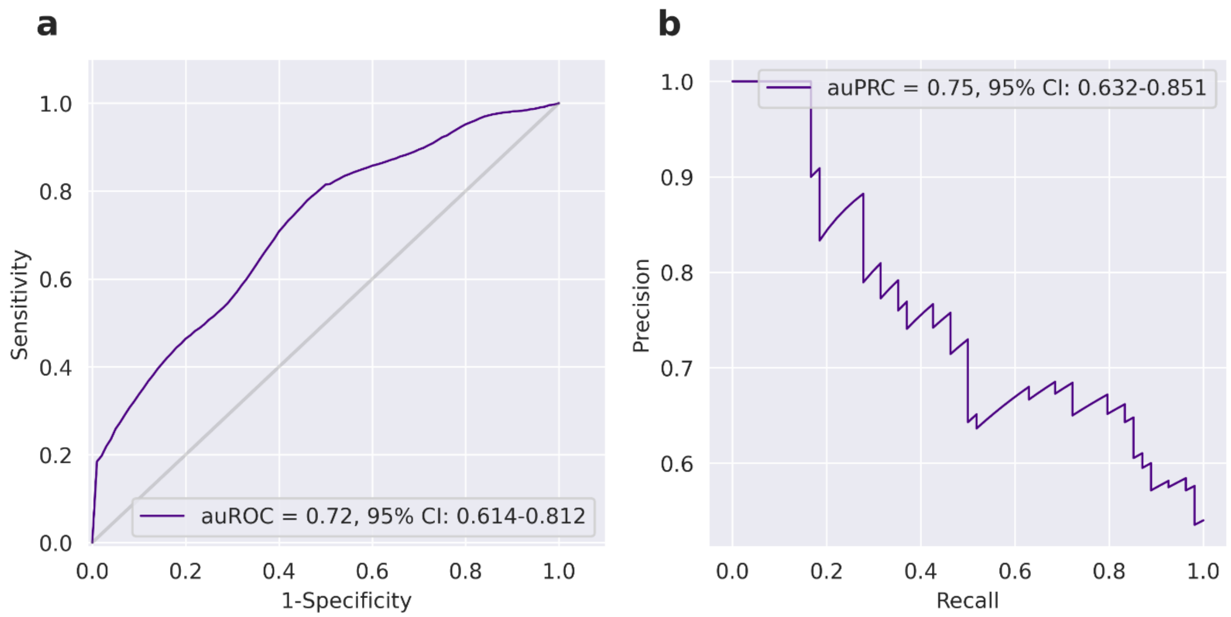


**Supplementary Figure 2. Performance of the inclusive model on ICU patients a**. Receiver-operating characteristics (ROC) curves. **b**. A plot of the precision (positive predictive value, PPV) against the recall (sensitivity) of the predictor for different thresholds.

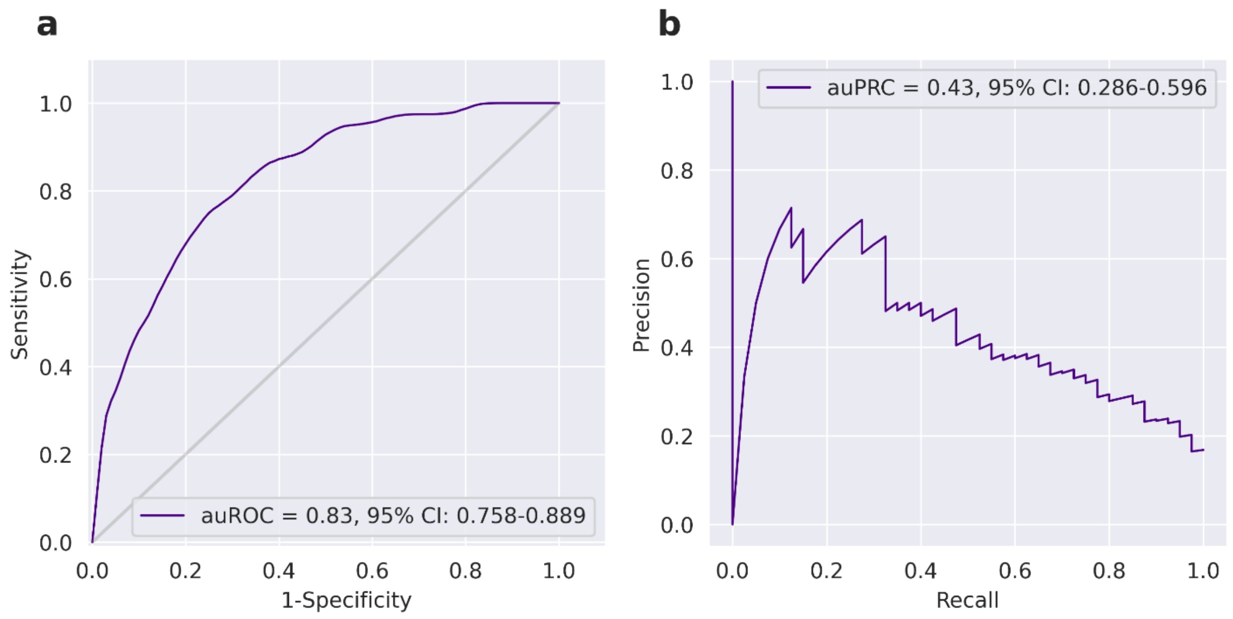


**Supplementary Figure 3. Performance of the inclusive model on ER patients a**. Receiver-operating characteristics (ROC) curves. **b**. A plot of the precision (positive predictive value, PPV) against the recall (sensitivity) of the predictor for different thresholds.
